# Cell graph neural networks enable the digital staging of tumor microenvironment and precise prediction of patient survival in gastric cancer

**DOI:** 10.1101/2021.09.01.21262086

**Authors:** Yanan Wang, Yu Guang Wang, Changyuan Hu, Ming Li, Yanan Fan, Nina Otter, Ikuan Sam, Hongquan Gou, Yiqun Hu, Terry Kwok, John Zalcberg, Alex Boussioutas, Roger J. Daly, Guido Montúfar, Pietro Liò, Dakang Xu, Geoffrey I. Webb, Jiangning Song

## Abstract

Gastric cancer is one of the deadliest cancers worldwide. Accurate prognosis is essential for effective clinical assessment and treatment. Spatial patterns in the tumor microenvironment (TME) are conceptually indicative of the staging and progression of gastric cancer patients. Using spatial patterns of the TME by integrating and transforming the multiplexed immunohistochemistry (mIHC) images as Cell-Graphs, we propose a novel graph neural network-based approach, termed *Cell-Graph Signature or CG*_*Signature*_, powered by artificial intelligence, for digital staging of TME and precise prediction of patient survival in gastric cancer. In this study, patient survival prediction is formulated as either a binary (*short-term* and *long-term*) or ternary (*short-term, medium-term*, and *long-term*) classification task. Extensive benchmarking experiments demonstrate that the *CG*_*Signature*_ achieves outstanding model performance, with Area Under the Receiver-Operating Characteristic curve (AUROC) of 0.960±0.01, and 0.771±0.024 to 0.904±0.012 for the binary- and ternary-classification, respectively. Moreover, Kaplan-Meier survival analysis indicates that the ‘digital-grade’ cancer staging produced by *CG*_*Signature*_ provides a remarkable capability in discriminating both binary and ternary classes with statistical significance (*p*-value < 0.0001), significantly outperforming the AJCC 8th edition Tumor-Node-Metastasis staging system. Using Cell-Graphs extracted from mIHC images, *CG*_*Signature*_ improves the assessment of the link between the TME spatial patterns and patient prognosis. Our study suggests the feasibility and benefits of such artificial intelligence-powered digital staging system in diagnostic pathology and precision oncology.

---

Gastric cancer (GC) accounted for 768,793 deaths in 2020, representing the fourth deadliest cancer globally^1^. The 5-year survival rate of GC is around 20%^2^. More accurate prognosis can greatly assist clinical decision-making, especially regarding which patients would benefit from aggressive treatment. The Tumor-Node-Metastasis (TNM) staging system^3^ is the most prevalent cancer staging system primarily used in hospitals and medical centers worldwide, which reflects the information of the primary tumor, affected lymph nodes, and metastasis. Many current treatment recommendations and guidelines are based on the TNM stages. However, significant differences in clinical outcomes have been observed in GC patients with the same TNM stage and similar treatment regimens^4–6^. These findings indicate the TNM staging system has limitations and accordingly, cannot be used to accurately predict prognosis of cancer patients. As such, new strategies that can provide more tailored staging information and improve prognosis predictions are highly desirable.

Recent years have seen numerous data-driven, machine learning-based studies of cancer prognosis. For instance, Yu *et al*. introduced prognosis prediction of lung adenocarcinoma and squamous cell carcinoma of stage I, and their model can distinguish the shorter-term survivors from longer-term survivors (*p <* 0.003 and *p* = 0.023)^7^. Mobadersany *et al*. presented survival convolutional neural network (SCNN), and their developed histology image-based SCNN reached comparable performance on astrocytomas of grade III and IV with histology grading or molecular subtyping^8^. In another study, Jiang *et al*. proposed the GC-SVM classifier as a powerful survival predictor using the data of immunomarkers and could predict the adjuvant chemotherapy benefit of gastric cancer patients with stages II and III^9^. Wulczyn *et al*. conducted a survival prediction study involving multiple cancers based on deep learning, and as a result, their model was capable of making significant survival predictions for five out of ten cancers and could effectively stratify cancer patients of stages II and III^10^. Jiang *et al*. developed a convolutional neural network-based classifier from H&E images to predict the prognosis of stage III colon cancer patients^11^. Dimitriou *et al*. introduced a K-nearest neighbor-based method to predict the mortality of stage II colorectal cancer patients using immunofluorescence images^12^. Although these prognosis prediction studies achieved promising performance using H&E staining histology or immunohistochemistry staining images, they were often restricted to specific subtypes or stages of the corresponding cancers. Moreover, these studies also did not consider any spatial information from the tumor microenvironment (TME).

Cell distribution in TME is not random but rather it is associated with the underlying functional state^13^. Therefore, the exploration of the TME of cancer samples would offer critical insights into the key spatial patterns associated with the growth, cancer progression, and thus patient prognosis^14^. Recent advent multiplexed immunohistochemistry (mIHC) staining technique enables systematic investigation of the TME^15,16^ and supports extraction of enriched spatial information from the TME, including the cell location, cell types, cell and nucleus morphological information, and related optical information^14,17^. Researchers have applied the mIHC technique to analyze the TME of pancreatic cancer and found that spatial distribution of cytotoxic T cells in proximity to cancer cells correlates with increased overall patient survival^14^. Barua *et al*. applied a statistical scoring based method, G-cross function, to measure the patterns of two different cell types, such as T-reg and CD8, and found that high infiltration of T-reg in the core tumor area is an independent predictor of worse overall survival (OS) in patients of non-small cell lung cancer^17^. However, these studies only considered the spatial features of limited cell types and only used handcrafted features. Therefore, comprehensive and quantitative methods that assess the relationships between spatial features descriptive of cell distribution and prognosis are currently lacking.

Inspired by the concept of the Cell-Graph^13,18^ and the success of graph neural networks (GNN)^19–21^, especially their applications to the analysis of biology data^22,23^, we hypothesize that intricate spatial distribution information of the TME is informative for the prediction of the OS of GC patients and a GNN model can effectively capitalize on the useful patterns generated by Cell-Graphs. To validate this hypothesis, we have developed a novel GNN-based approach for predicting the prognosis of GC patients using Cell-Graph data, which we call the *Cell-Graph Signature* or *CG*_*Signature*_. The overall workflow is illustrated in Figure 1 and Figure S1. In this study, we formulate prognosis prediction as a classification problem by predicting the patient’s survival time interval rather than a continuous time frame or a risk score and develop a workflow to perform the following three-fold tasks. Firstly, it extracts the comprehensive spatial and morphological information from mIHC images. Secondly, it further uses the extracted spatial information to stratify patients into either binary (*short-term* and *long-term*) or ternary (*short-term, medium-term*, and *long-term*) classes. Finally, it conducts the Kaplan-Meier survival analysis to verify the clinical significance of the *CG*_*Signature*_.

**Figure 1.**
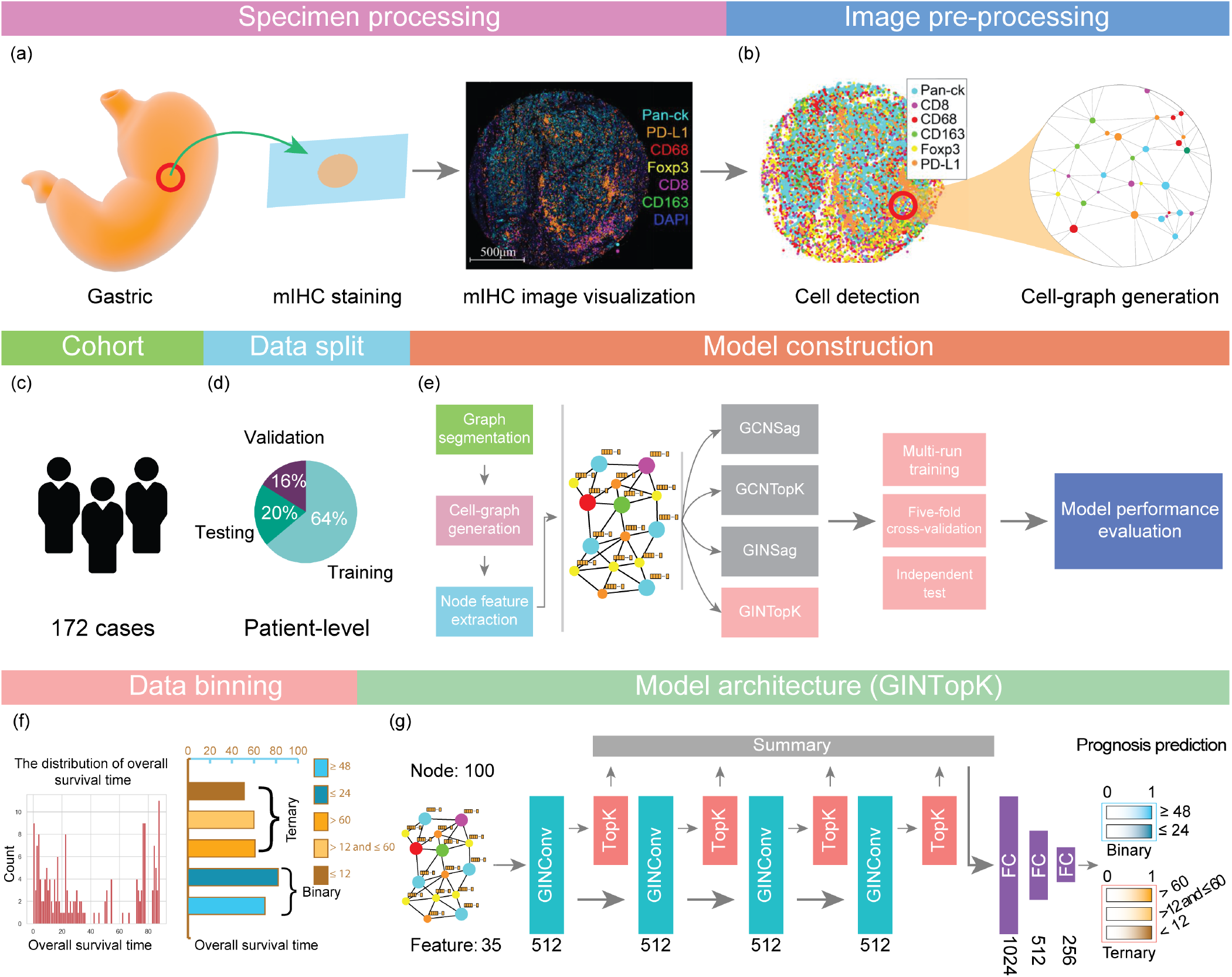
An overall workflow of graph neural network-based prognosis prediction using Cell-Graphs. **(a)** Specimen processing: The tumor tissues were extracted from gastric cancer, and stained with seven different biomarkers including DAPI, Pan-CK, CD8, CD68, CD163, Foxp3, and PD-L1. **(b)** Image pre-processing: sub-sampling and cell-graph construction were conducted for image pre-processing. **(c)** An illustration for the cohort, 172 gastric cancer patients were collected. **(d)** Data split. The training, validation and testing datasets were split with the percentages of 64%, 16%, and 20%, respectively. **(e)** Model construction: four different GNN model architectures, including GCNSag, GCNTopK, GINSag, and GINTopK, were constructed and compared. Multi-run model training, five-fold cross-validation, and independent test were conducted to evaluate the performance of the constructed GNN models. **(f)** Data binning: overall survival time ranged from 0 to 88 months, and two data binning strategies were applied to generate binary- and ternary-class datasets. **(g)** Model architecture: The four models shared the same architecture but employed different types of convolutional unit and pooling layer, which consists of four consecutive convolutional layer and pooling layer blocks, followed by a summary layer and three fully-connected layers, prior to the generation of the final classification outcome. Architecture of the best-performing GINTopK model is illustrated herein, which outperformed the other three model architectures and also achieved the best performance on the test dataset. The corresponding number of hidden layers or feature dimensions are indicated at the bottom of each box. Here, FC stands for “fully connected layer”.

*CG*_*Signature*_ represents a powerful survival predictor under comprehensive and extensive benchmarking tests of gastric cancer across all subtypes and stages. Specifically, *CG*_*Signature*_ can effectively stratify short-term, medium-term, and long-term GC survivors at the early diagnosis stage, and achieved the area under the receiver-operating characteristic curve (AU-ROC) values of 0.960 ± 0.01 in terms of binary classification, and 0.771 ± 0.024 to 0.904 ± 0.012 in terms of ternary classification, respectively. In the follow-up survival analysis, *CG*_*Signature*_ outperformed the AJCC 8th edition TNM staging system on the testing cohort in terms of the Harrell’s Concordance-Index^24^, Hazard Ratio (HR), and *p*-value.

## Results

### Clinical characteristics and data-binning of the patient cohort

We collected the data of 172 gastric cancer patients from Shanghai Ruijin Hospital, affiliated with the School of Medicine, Shanghai Jiao Tong University. The clinical characteristics of this cohort are illustrated in Table S1. This cohort contains 124 males, 47 females, and one case without gender information. With respect to the survival status, 113 cases were recorded as “death” while 59 patients as “live”. The statistical summary of their TNM (the AJCC 8th edition) stages are provided in Table S1. In particular, the patient numbers of the TNM stages of I, II, III, and IV are 14, 52, 95, and 3, respectively. The OS time of the cohort ranges from 0 to 88 months. Two data-binning strategies were applied to segment the patient OS into binary- or ternary-class datasets. More specifically, patients with OS time shorter than 24 months and longer than 48 months were categorized as short-term, and long-term in the binary-class dataset. The patients whose OS time is between 24 months and 48 months were removed from the training dataset but used in subsequent survival analyses. More details can be found in the section of **Survival analysis and performance comparison with the TNM staging system**. In the ternary-class dataset, patients were classified into short-term, medium-term, and long-term classes, using the thresholds of 12 and 60 months. Here, the data-binning thresholds were chosen to take into account the relative class balance and clinical importance. We did not optimize the data binning threshold, which, however, can be conducted when more data becomes available. Model training and subsequent analysis were performed using these two datasets.

### Workflow overview

Figure 1 illustrates an overall workflow and the model architecture of the proposed *CG*_*Signature*_ approach. As shown in Figure 1a, the mIHC technique was used to stain the GC tissue samples. Specifically, the nuclear counterstain, DAPI, was used for cell nuclei staining, and six antibodies of Pan-CK, CD8, CD68, CD163, Foxp3, and PD-L1 were used as annotation indicators for six different types of cells. After digitalization, cell locations, types, and related optical and morphological features were extracted using the digital pathology software. After this procedure, we obtained the CSV files in which each row corresponds to each cell with the node features shown in Table 3. Based on these CSV files as the input, we developed a workflow (details can be seen in Algorithm 1) to process the raw data and build the GNN-based model to predict the patient OS interval using the features extracted from mIHC images.

The key steps of the workflow are as follows: **(1) Image pre-processing**: Sub-sampling and Cell-Graph generation were performed at this step. Specifically, each mIHC image was firstly segmented into multiple non-overlapping regions with no more than 100 cells. For each region, we built a graph where each cell was represented as a node and the reciprocal of the Euclidean distance of each cell-cell pair was used to establish edges between them with the distance of less than 20 *µm*. Detailed information can be found in the section “Cell-Graph construction” of Methods. Then, we extracted a total of 35 features (as shown in Table 3) for each cell as the node attributes, including 5 optical features for each biomarker and 5 morphological features for each cell. Such generated cell-based graph is referred to as Cell-Graph^13,18^. There are approximately 90 Cell-Graphs constructed for each mIHC image (for each patient). Cell-Graphs originated from the same mIHC image share the same label with the corresponding patient. **(2) Data split**: After Cell-Graph construction, the whole dataset was partitioned into the training, validation, and test sets with the ratio of 0.64 : 0.16 : 0.20 at the patient level. In addition, we also generated the files for performing five-fold cross-validation by generating five non-overlapping training-validation subsets and evaluating the model performance on these 5-fold subsets. **(3) Hyper-parameter optimization**: We utilized the Hyperopt toolkit^25^from the Ray software package^26^ to tune the hyperparameters of GNN models. The optimized hyperparameters were then used for the follow-up model training and performance evaluation. **(4) Model performance evaluation and data visualization**: To comprehensively assess the capability and reliability of our GNN model, we evaluate model performance using multi-run model training, five-fold cross-validation, and independent test. The test results were visualized by generating the receiver-operating characteristic (ROC) curves, confusion matrix, and boxplots of Accuracy, F1-Score, and Matthews Correlation Coefficient (MCC). Performance metrics are defined in Section “Metrics of model performance evaluation” in the Supplementary material.

### Performance benchmarking of different GNN models for prognosis prediction

We constructed four different types of GNN models and examined their performance for predicting the OS of gastric cancer patients, including GINTopK, GINSAG, GCNTopK and GCNSAG. Here, GIN^21^ and GCN^27^ are two graph convolution computational units (differences can be seen in Figure S2), whereas TopKPooling^20,28,29^ and SAGPooling^29,30^ are two graph pooling computational units. The graph convolutional and pooling layers are the core components of the GNN architecture. Five-fold cross-validation was conducted to assess the model of each GNN model on both binary- and ternary-classification tasks. The results are averaged on ten repetitions of five-fold cross-validation for GINTopK on binary classification (as shown in Figure S3) to circumvent the randomness of the model during training. In this procedure, Accuracy, F1-Score, MCC, and AUROC were calculated to evaluate the performance. Figure 2a illustrates the performance results of binary classification on five-fold cross-validation. As we observe, the median values of both Accuracy and F1-score for the four GNNs ranged from 0.83 to 0.92, while the median values of MCC ranged from 0.66 to 0.84, respectively. Figure 2b shows the performance results of ternary classification on five-fold cross-validation. We can see that the ternary-class classification models achieved the median values of Accuracy ranging from 0.76 to 0.82, F1-score from 0.64 to 0.72, and MCC from 0.46 to 0.5, respectively. According to the results shown in Figures 2a and 2b, GINTopK slightly out-performed the other three GNN models on both binary- and ternary-classifications. Therefore, GINTopK was selected as the best-performing GNN model and employed for subsequent performance benchmarking and survival analysis.

**Figure 2.**
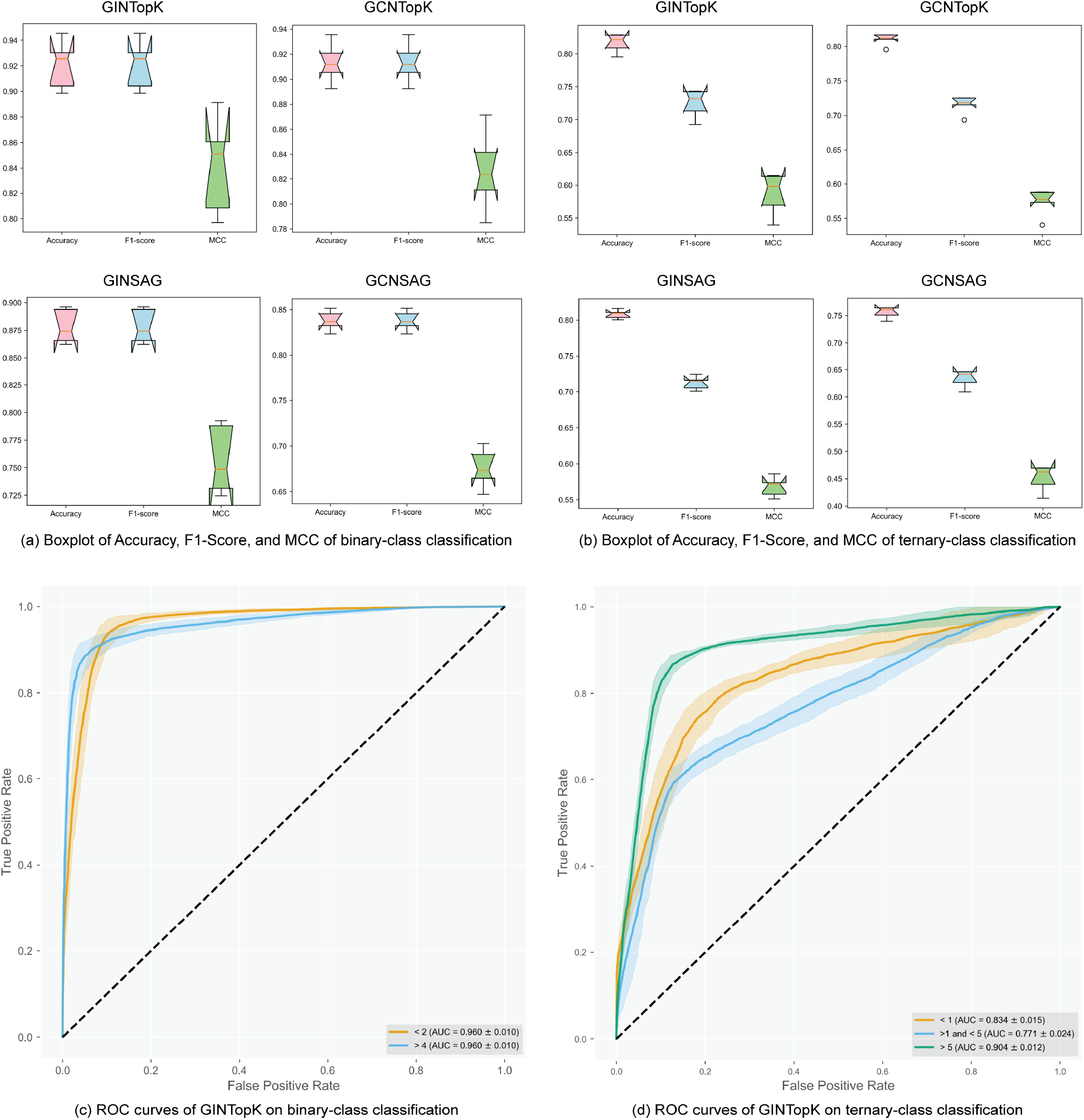
Model performance of four GNNs on five-fold cross-validation. (a) and (b) show the Boxplots of performance metrics of Accuracy, F1-score, and MCC on five-fold cross-validation. (c) and (d) illustrate the ROCs of GINTopK binary- and ternary-models on five-fold cross-validation.

ROC curves of GINTopK on the binary- and ternary-classification tasks are illustrated in Figure 2c and Figure 2d, respectively. The binary-class GINTopK model achieved the AUROC value of 0.96 ± 0.01 on five-fold cross-validation. In contrast, the ternary-class GINTopK classifier reached the AU-ROC values of 0.834 ± 0.015, 0.771 ± 0.024, and 0.904 ± 0.012 for *short-term* (<12 months), *medium-term* (>12 and <60 months), and *long-term* (>60 months) on five-fold cross-validation, respectively (Figure 2d). Moreover, the performance results of binary-class GINTopK model on ten repetitions of five-fold cross-validation are displayed in Figure S3. We can see that the median values of both Accuracy and F1-score were within the range of 0.90-0.93 (MCC values ranged from 0.80 to 0.86), thereby suggesting the stability of our proposed GINTopK model.

In Figure 3, the performance results of the GINTopK model on the independent test are visualized using ROC curves and confusion matrix. It can be seen that the model achieved similar performance with that on five-fold cross-validation in terms of AUROC values on both binary- and ternary-classification tasks. In terms of the confusion matrix, 96% and 89% of the short-term and long-term patients could be accurately predicted using the binary-classification model. The true positive percentages of ternary-class model were 81%, 59%, and 85%, corresponding to the short-term, medium-term, and long-term classes (Figure 3).

**Figure 3.**
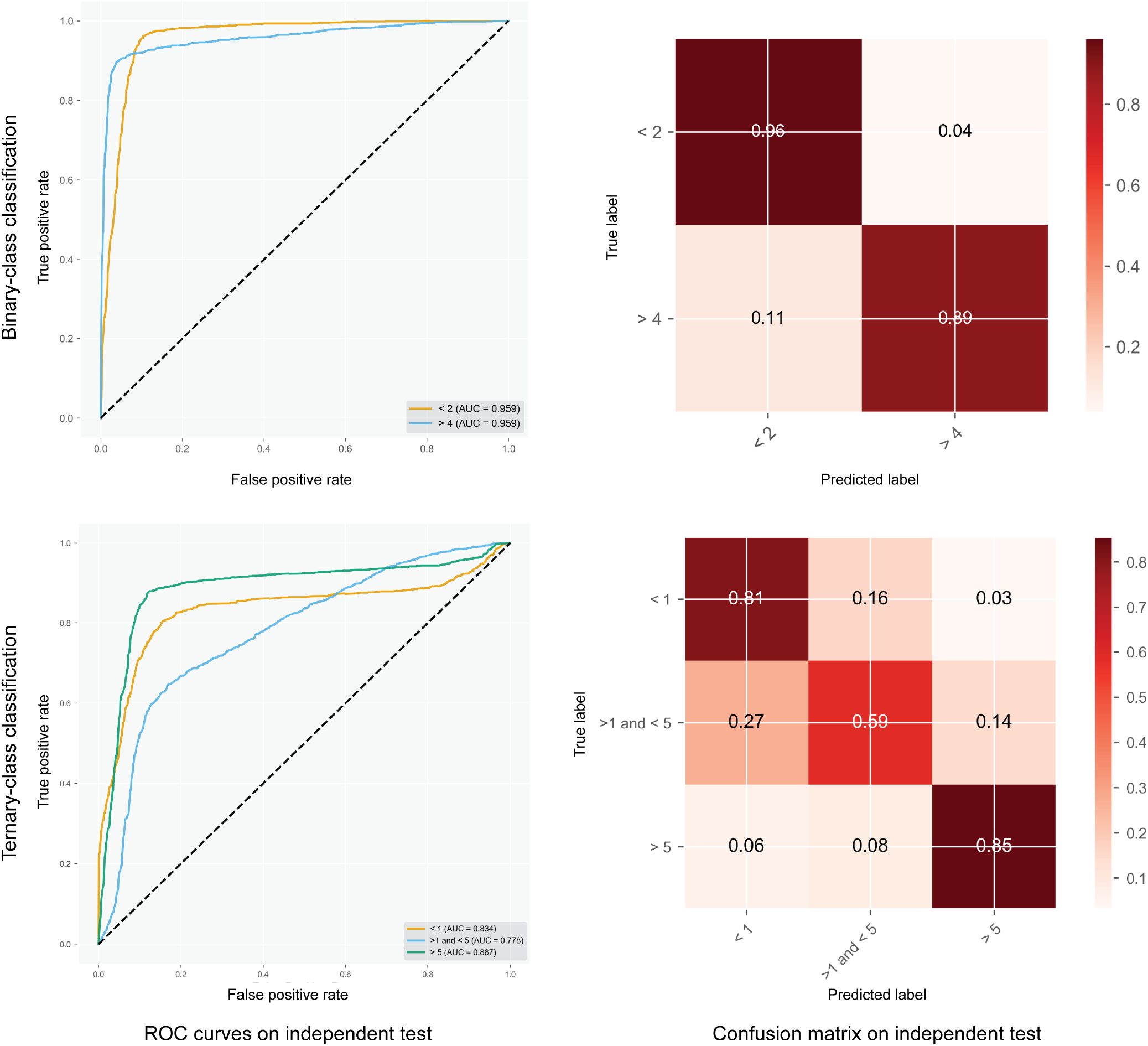
Performance assessment of the GINTopK model in terms of ROC curves and confusion matrix on the independent test. The left column shows the ROC curves of binary- and ternary-classification, while the right column displays the confusion matrix of the model predictions on the binary- and ternary-classification tasks.

Taken together, outstanding performance of the GINTopK model on both cross-validation and independent test indicate that our proposed GNN approach is capable of effectively capturing the underlying prognostic patterns from the well-constructed Cell-Graphs. The captured prognostic patterns by GNN model are characteristic of the spatial information of cell locations and types of the TME, which incorporates more potentially informative features than the TNM staging system.

### Ablation studies and prognostic value of different types of cell features

To examine the effect of node features of different cell types on model performance, we further performed ablation studies to assess the contribution of features to the binary- and ternary-classification performance by removing each type of features in an iterative manner. Thirty-five node features of seven types were used in this study, including DAPI, Pan-CK, CD8, CD68, Foxp3, PD-L1, and morphological features. We first evaluated the performance of the GNN model trained using all these features, and then, evaluated the performance of the models trained using the remaining features after removing each type of features from the all-feature set in turn. For each iteration, we trained the models five times with random initialization of the weights using the same dataset and calculated the mean and standard deviation of Accuracy. The results are shown in Table 2, where the feature contribution was measured by the accuracy change compared with that of the all-feature model. Note that when a type of feature is removed, an accuracy increase means that including the feature type reduced accuracy, and an accuracy decrease means that the feature type played an important role in attaining the all feature accuracy.

According to Table 2, the variant models trained using these feature subsets and all-feature set achieved comparable Accuracy values in both binary and ternary classifications. In the binary classification, the DAPI features and morphological features made more important contributions to the model performance compared with other types of features (e.g. the Accuracy dropped by 0.035 and 0.025, respectively), which reflect the nucleus differences of optical and morphology of the TME. Thus, inclusion of these two types of features helped to better distinguish the long-term from short-term patients. In the case of ternary classification, we can see that the GNN models trained without the of DAPI and morphology features achieved the lowest Accuracy, which is consistent with the observation in the binary classification.

### Survival analysis and performance comparison with the TNM staging system

To further investigate the prognostic values and clinical importance of the predictions produced by *CG*_*Signature*_, we conducted the Kaplan-Meier survival analysis using the patient-level results of both binary- and ternary-classifications. For each patient, we first collected the predicted results of all the subsampled Cell-Graphs. Next, we calculated the class percentages of these predictions, and took the class with the maximum percentage as the final patient-level prediction of the corresponding patient. Using these patient-level predicted results (‘digital-grade’) of binary-classification (with predicted class labels of *CG*_*Signature*_ = 0 and *CG*_*Signature*_ = 1) and ternary-classification (with predicted class labels of *CG*_*Signature*_ = 0, *CG*_*Signature*_ = 1, and *CG*_*Signature*_ = 2), we conducted the survival analysis and plotted their Kaplan-Meier curves, shown in Figure 4. More specifically, when using the binary-class predictions, the median survival time of patient test cohorts predicted as *CG*_*Signature*_ = 0 and *CG*_*Signature*_ = 1 were about 18 months and 42 months, respectively. The hazard ratio was 0.217 (95% CI: 0.108 – 0.438), the C-Index was 0.699 (95% CI: 0.637 – 0.762), and the *p*-value was less than 0.0001, indicating that *CG*_*Signature*_ has statistically significant prognostic power in separating the two groups of patient cohorts. When using the ternary-class predictions, the median survival time of patient cohorts predicted as *CG*_*Signature*_ = 0 and *CG*_*Signature*_ = 1 were around 7 months and 28 months, respectively. The endpoint survival rate of *CG*_*Signature*_ = 2 was approximately 92.3% (Figure 4b). The hazard ratio and C-Index were 0.204 (95% CI: 0.107 – 0.389) and 0.823 (95% CI: 0.748 – 0.899), respectively, with the *p*-value less than 0.0001.

**Figure 4.**
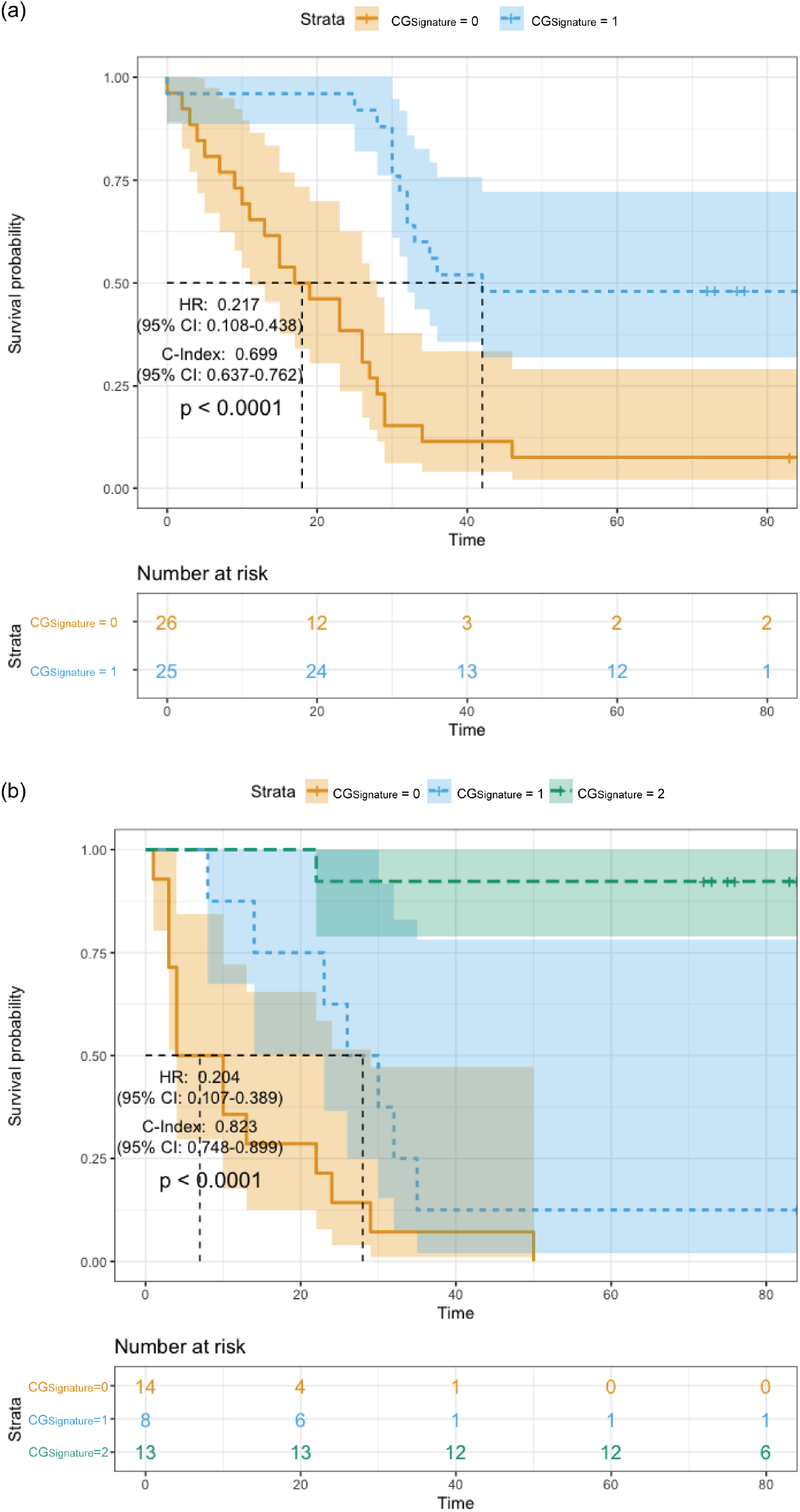
Kaplan-Meier survival analysis of patient overall survival based on the ‘digital grade’ (patient-level predictions) producted by *CG*_*Signature*_ in terms of **(a)** binary- and **(b)** ternary-classification. As can be seen from Figure 4, Kaplan-Meier survival analysis demonstrates that the ‘digital grade’ cancer staging produced by *CG*_*Signature*_ provides a remarkable capability in discriminating both binary (short-term, long-term) and ternary (short-term, medium-term, and long-term) classes with C-Index (Binary: 0.699 (95% CI: 0.637 – 0.762), Ternary: 0.823 (95% CI: 0.748 – 0.899)), Hazard Ratio (Binary: 0.217 (95% CI: 0.108 – 0.438), Ternary: 0.204 (95% CI: 0.107 – 0.389)), and the *p*-values < 0.0001.

We further compared the patient survival analysis based on predictions of *CG*_*Signature*_ with the AJCC 8th edition TNM staging system and showed the results in Table 1. In the TNM staging system, there were eight groups of *I*_*A*_, *I*_*B*_, *II*_*A*_, *II*_*B*_, *III*_*A*_, *III*_*B*_, *III*_*C*_, *IV*_*A*_, and *IV*_*B*_. As no patients of stage *IV* were included in the binary-class test cohort and only one patient of stage *IV* was included in ternary-class test cohort, we excluded the patients of stage *IV* and those without OS information. Finally, 51 patients (including 20 uncategorized patients) and 35 patients were retained for binary- and ternary-class survival analysis, respectively. The detailed statistical information of the testing cohorts can be found in Table S2.

**Table 1.** Overall Kaplan-Meier survival analysis based on the predictions of binary- and ternary-classification by *CG*_*Signature*_. The classification results were compared with Harrell’s Concordance-Index (C-Index), Hazard Ratio (HR), and *p*-value. For the convenience of survival analysis comparison, the variables of TNM stages were regrouped into TNM-2 (I+II vs. III), TNM-3 (I vs. II vs. III), and TNM-6 (I, IIA, IIB, IIIA, IIIB, IIIC), while “*CG*_*Signature*_+TNM-2” denotes a four-class variable by combining the classes of TNM-2 and binary-class *CG*_*Signature*_.

| | Variable | C-Index (95% CI) | HR (95% CI) | $p$-value |
| --- | --- | --- | --- | --- |
| Binary-class test cohort | TNM-2 (I+II vs. III) | 0.659 (0.577 – 0.740) | 5.276 (2.147 – 12.966) | <0.0001 |
|  | TNM-6 (I, IIA, IIB, IIIA, IIIB, IIIC) | 0.714 (0.623 – 0.805) | 1.873 (1.388 – 2.529) | 0.00081 |
| | $CG_{Signature}$ (Low vs. High) | 0.699 (0.637 – 0.762) | 0.217 (0.108 – 0.438) | <0.0001 |
| | $CG_{Signature}$ + TNM-2 | **0.740** (0.661 – 0.819) | **2.412** (1.650 – 3.525) | <0.0001 |
| Ternary-class test cohort | TNM-3 (I vs. II vs. III) | 0.632 (0.510 – 0.753) | 3.169 (1.335 – 7.522) | 0.019 |
|  | TNM-6 (I, IIA, IIB, IIIA, IIIB, IIIC) | 0.681 (0.535 – 0.827) | 1.708 (1.212 – 2.407) | 0.028 |
| | $CG_{Signature}$ (Low vs. Medium vs. High) | **0.823** (0.748 – 0.899) | **0.204** (0.107 – 0.389) | <0.0001 |

To make a fair comparison, three specific criteria were adopted to aggregate the TNM stages into TNM-2 (*I, II* vs. *III*), TNM-3 (*I* vs. *II* vs. *III*), and TNM-6 (*I* vs. *II*_*A*_ vs. *II*_*B*_vs. *III*_*A*_ vs. *III*_*B*_ vs. *III*_*C*_). The survival analysis results are provided in Table 1 and Figures S4-S9. According to Table 1 and Figure S4, the C-Index of the binary-class *CG*_*Signature*_ was 0.699 (*p*-value *<* 0.0001), outperforming TNM-2 with an increase of 0.04. We further combined the TNM-2 with binary- class *CG*_*Signature*_ for survival analysis (Figure S6), which achieved the highest C-Index of 0.748 (*p*-value *<* 0.0001), which was higher than TNM-6 by 0.034 (Figure S5). In ternary-class survival analysis, we compared the results of TNM-3, TNM-6, and the ternary-class *CG*_*Signature*_. More specifically, C-Index of the ternary *CG*_*Signature*_ was 0.823 (*p*-value *<* 0.0001, Figure 4b), which was superior to the TNM-3 (Figure S8) and TNM-6 (Figure S9) with an increase of 0.191 and 0.142, respectively. These results demonstrate the *CG*_*Signature*_ is capable of discriminating and stratifying gastric cancer patients into groups of different prognosis better than the TNM staging system. Moreover, we note that the prognostic power can be even further enhanced by integrating − the *CG*_*Signature*_ predictions and the TNM stages for survival analysis, such as the *CG*_*Signature*_ + TNM − 2 in Table 1 and Figure S6.

To summarize, by combining the spatial information from the mIHC images, *CG*_*Signature*_ has demonstrated outstanding performance in survival analysis, and achieved a better or at least comparable performance when comparing with the TNM staging system. The results suggest that effective prognostic features can indeed be captured by *CG*_*Signature*_, which suggests a powerful method complementary to the current TNM staging system.

### Framelet decomposition for cell-graph

To examine the capacity of Cell-Graph to capture useful spatial features from mIHC images, we conducted a framelet decomposition on the whole mIHC images. The framelet transforms (including framelet decomposition and reconstruction) have proved an important tool for distilling multi-resolution information in low-pass and high-passes from the graph data^31–35^.

We extracted low-pass and high-pass information of six types of features, corresponding to six different biomarkers DAPI, PAN-CK, CD8, CD68, FOXP3, and PD-L1. Tables S3-S11 show the low-pass and high-pass coefficients of the framelet decomposition on mIHC images of short-term, medium-term and long-term survivors on the entire mIHC images. For the selected samples, no significant differences were observed from the low-pass channel. However, major differences can be observed from the high-pass channel on the selected samples. More specifically, remarkable signal differences can be seen from the high-pass channel-1 and channel-2 in terms of the features of Cell Area and Nucleus Perimeter (summarized in Table S6-S11). These differences highlight the important prognostic value of cell morphological information of the TME, which is consistent with the prognostic value of different types of cell features.

## Discussion

In this study, we developed the first GNN-based approach, Cell-Graph Signature (*CG*_*Signature*_), which is capable of predicting the prognosis of gastric cancer patients from Cell-Graphs extracted from mIHC images. Extensive benchmarking tests on multi-run model training, 5-fold cross-validation, and independent test demonstrate that *CG*_*Signature*_ can accurately predict the prognosis on both binary- and ternary-class classification tasks. We designed and compared the performance of four different GNN architectures, including GINSag, GCNTopK, GCNSag, and GINTopK. As a result, GINTopK achieved the best performance when compared with the other three GNN architectures (GINSag, GCNTopK, and GCNSag) on the same datasets. Feature ablation studies showed that the nucleus optical feature (DAPI) and cell morphological features are essential node features for and contributed most to the prognosis prediction, which indicate the potential pivotal roles of nuclear and cell morphology in gastric cancer progression. In survival analysis, *CG*_*Signature*_ clearly outperformed the AJCC 8th TNM staging system in terms of C-Index (0.823, 95% CI: 0.748-0.899) using the ternary-classification model. In particular, we notice that *CG*_*Signature*_ achieved better or comparable performance with the TNM staging system when using the binary-classification model. These results of survival analysis indicate that *CG*_*Signature*_ provides more prognostic power than the existing TNM staging system and can help pinpoint patients who may benefit from more tailored and personalized therapy. Moreover, wavelet decomposition results suggest that Cell-Graphs can indeed capture certain important spatial features informative for classifying patient survival. Although many previous studies of prognosis prediction also achieved promising results, the majority of such studies were only limited to a specific subtype or stage of cancers. Nevertheless, in this study, we show that the proposed *CG*_*Signature*_ method is applicable to gastric cancer patients of all subtypes across all TNM stages. Moreover, *CG*_*Signature*_ achieved a better performance when stratifying test patient cohorts into different groups of prognosis, which has proven a powerful prognostic predictor for gastric cancer.

One caveat of the current study is that we could only obtain a limited size of the mIHC image data, and accordingly, the performance of the *CG*_*Signature*_ was only benchmarked on the limited size of the gastric cancer patient dataset. Thus, in future studies, it would be important to evaluate the performance of graph neural networks based on Cell-Graph data from mIHC images in much larger and/or multi-centre patient cohorts, as well as additional tumor types (in addition to gastric cancer), when more data become available. Exploration of the prognostic value of the *CG*_*Signature*_ method on datasets of other cancer types would surely be needed to verify its utility and capability. Additionally, future extension of the capability of *CG*_*Signature*_ by using whole-slide images and other biomarkers in mIHC/mIF staining, for example, holds great potential for a more comprehensive analysis of the tumor microenvironment^15^; this will in turn serve to better inform the training of more accurate GNN models. The continuing development of cutting-edge, robust, and broadly-applicable Cell Graph-based biomarker discovery algorithms is valuable and desirable to better inform and transform the medical care of cancer patients.

## Methods

### Dataset

The gastric cancer samples were collected and stained with mIHC technique and prepared as two batches of tissue microarray^36^, in which all the samples were arranged in the matrix configuration. Then the two tissue microarrays were scanned by digital microscope (brand: Vectra Polaris) under the magnification of 40X with each pixel represents 0.5 *µm*. Totally, 181 mIHC images of cancer tissues were curated as the initial datasets. After excluding patients whose follow-up data were not available, 172 mIHC images were retained and used for model training and benchmarking. The overall survival time of the patients ranges from 0 to 88 months, as shown in Figure 1f. Detailed clinical characteristics and statistical summary of the cohort are provided in Table S1. Fifty-nine patients were still alive at the time of the last follow-up. All the images were stained using multiplexed immunohistochemistry of seven colors and reagents to identify the specific cell types. In this study, cells were stained with antibodies of Pan-CK, Foxp3, CD8, PD-L1, CD68, CD163, and DAPI. The dataset was randomly partitioned into the training, validation, and test subsets with the ratios of 0.64, 0.16, and 0.20 at the patient level. In addition, datasets for five-fold cross-validation were also prepared.

### Label generation

In this study, the survival prediction was formulated as a classification problem in the form of either binary- or ternary-classification. To explore the prognostic value of the Cell-Graphs extracted from the gastric cancer TME, the survival time of the cohort was categorized into two and three classes, and used as labels for training binary- and ternary-classification models based on graph neural networks. In binary-classification, 82 patients with survival time of less than 24 months were annotated as short-term while 70 patients with survival time of longer than 48 months were annotated as long-term. 20 patients with survival time between 24 and 48 months were removed from the training dataset, and denoted as uncategorized patients. For the ternary-classification, 12 months and 50 months were respectively used as the thresholds to divide patients into short-, medium-, and long-term, with the corresponding patient numbers of 51, 60, and 61, respectively.

### Cell segmentation

After digitization, the mIHC images were pre-processed using the pathology software HALO (Indica Labs) for cell segmentation and feature extraction. The extracted information was subsequently saved as a CSV file in which each row represents the features of a cell (as shown in Table 3), including the cell locations, optical features of stained cells, and morphology features. Thirty-five of such features were selected as the node features for each cell. Detailed information can be found in the **Node attributes** section.

**Table 2.**
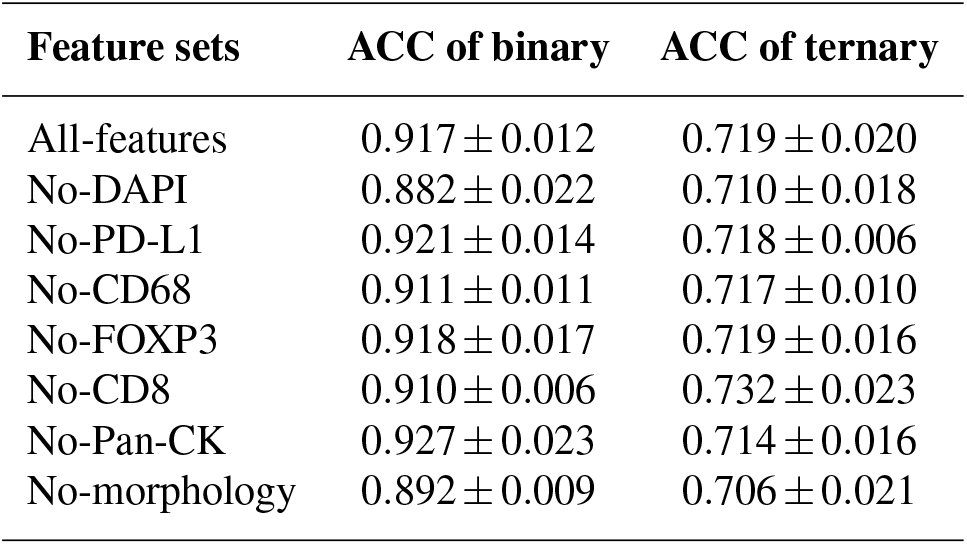
Ablation studies of the major types of features used by the GNN models in both binary and ternary-classification. The relative importance and contribution of the features was measured by the accuracy change compared with that of the all-feature model.

**Table 3.**
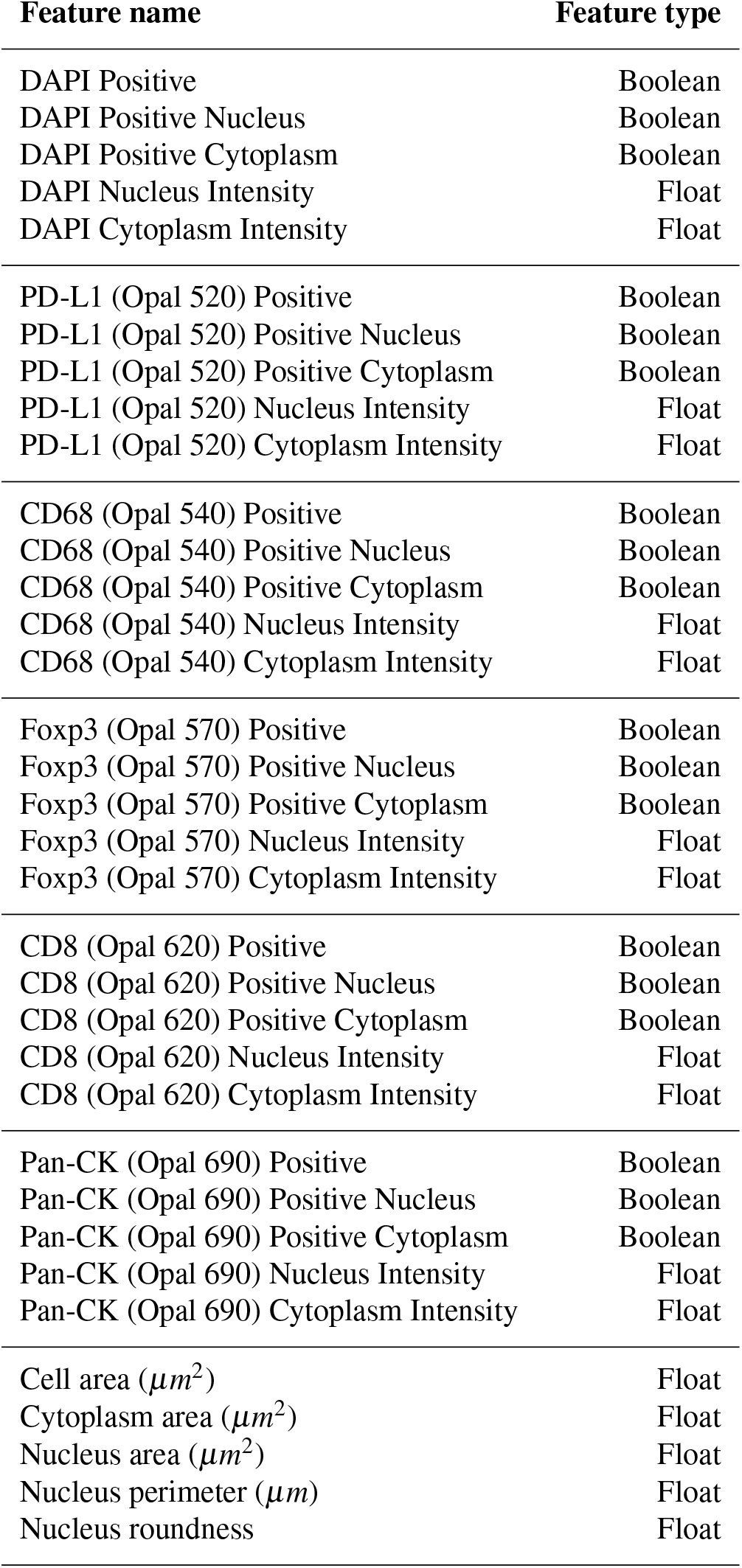
The list of node attributes and their variable types. Each type of features are comprised of three Boolean variables and two float variables. These Boolean variables were identified by the pathology software based on the float values of Nucleus Intensity and Cytoplasm Intensity of each biomarker. Moreover, five different morphology features were extracted as the node attributes.

| Feature name | Feature type |
| --- | --- |
| DAPI Positive | Boolean |
| DAPI Positive Nucleus | Boolean |
| DAPI Positive Cytoplasm | Boolean |
| DAPI Nucleus Intensity | Float |
| DAPI Cytoplasm Intensity | Float |
| PD-L1 (Opal 520) Positive | Boolean |
| PD-L1 (Opal 520) Positive Nucleus | Boolean |
| PD-L1 (Opal 520) Positive Cytoplasm | Boolean |
| PD-L1 (Opal 520) Nucleus Intensity | Float |
| PD-L1 (Opal 520) Cytoplasm Intensity | Float |
| CD68 (Opal 540) Positive | Boolean |
| CD68 (Opal 540) Positive Nucleus | Boolean |
| CD68 (Opal 540) Positive Cytoplasm | Boolean |
| CD68 (Opal 540) Nucleus Intensity | Float |
| CD68 (Opal 540) Cytoplasm Intensity | Float |
| Foxp3 (Opal 570) Positive | Boolean |
| Foxp3 (Opal 570) Positive Nucleus | Boolean |
| Foxp3 (Opal 570) Positive Cytoplasm | Boolean |
| Foxp3 (Opal 570) Nucleus Intensity | Float |
| Foxp3 (Opal 570) Cytoplasm Intensity | Float |
| CD8 (Opal 620) Positive | Boolean |
| CD8 (Opal 620) Positive Nucleus | Boolean |
| CD8 (Opal 620) Positive Cytoplasm | Boolean |
| CD8 (Opal 620) Nucleus Intensity | Float |
| CD8 (Opal 620) Cytoplasm Intensity | Float |
| Pan-CK (Opal 690) Positive | Boolean |
| Pan-CK (Opal 690) Positive Nucleus | Boolean |
| Pan-CK (Opal 690) Positive Cytoplasm | Boolean |
| Pan-CK (Opal 690) Nucleus Intensity | Float |
| Pan-CK (Opal 690) Cytoplasm Intensity | Float |
| Cell area ($\mu m^2$) | Float |
| Cytoplasm area ($\mu m^2$) | Float |
| Nucleus area ($\mu m^2$) | Float |
| Nucleus perimeter ($\mu m$) | Float |
| Nucleus roundness | Float |

### Sub-sampling

Each mIHC staining image contains around 7, 000 ∼ 13, 000 cells. In particular, we conducted the sub-sampling when generating the Cell-Graphs. By treating each cell as a node in the Cell-Graph, we limited the graph size with no more than 100 nodes. A non-overlap sliding window was then applied to extract the local regions that contained approximately 100 cells from the mIHC images. As a result, we obtained 16951 Cell-Graphs, which would be used for GNN model training and testing. The extracted Cell-Graphs from one mIHC image were annotated with the same label as that of the corresponding mIHC image. The performance of the GNN models was firstly assessed at the Cell-Graph level; After that, the prediction outputs of all Cell-Graphs were aggregated to generate the votes for the final prediction outcome at the patient level.

### Cell-Graph construction

According to the previous study on the TME^17^, we assumed that the maximum effective distance was 20 *µ*m between immune and tumor cells, which is equivalent to 40 pixels in the magnification of this study. We calculated the Euclidean distance between any pair of cells, and used this distance to define the edge weight between them according to the equations (1) and (2) shown below.

For the *i*th and *j*th cells with Cartesian coordinates (*x*_*i*_, *y*_*i*_) and (*x* _*j*_, *y* _*j*_) (which use pixel as the unit) in the same mIHC image, their Euclidean distance can be calculated as follows:

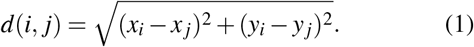

The weight between the *i*th and *j*th cells is assigned as follows:

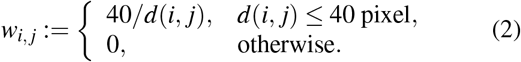

where 0 denotes that there is no interaction between the cell *i* and *j*. After sub-sampling, a number of Cell-Graphs (up to 100 nodes) were extracted and annotated, with the weight (2) of the edge between a given pair of cells.

### Node attributes

Graph neural network (GNN) is a powerful deep learning approach which can efficiently extract features from graph-structured data. In the present study, we focused on distilling five morphology features and 30 optical features generated by six staining biomarkers as the attributes of the node for each cell, including DAPI, PAN-CK, CD8, CD68, FOXP3, and PD-L1. The five morphology features include cell area, cytoplasm area, nucleus area, nucleus perimeter, and nucleus roundness. The optical features of each biomarker are comprised of positive, positive nucleus, positive cytoplasm, nucleus intensity, and cytoplasm intensity. As a result, a total of 35 features were extracted for each cell. These features indicates the area, shape, location and healthiness of the underlying cell. The detailed list of the features and their data types are listed in Table 3. All the features were linearly normalized to the range of [0, 1] prior to training the GNN models.

### Architecture of the designed graph neural networks

Graph-structured data are usually represented in the form of (*x*_*i*_, *A*_*i*_), where *x*_*i*_ denotes the feature of the node for the *i*th graph sample while *A*_*i*_ represents its adjacency matrix. A GNN has the similar network architecture to that of the traditional convolutional neural network. To address the classification task in this study, we designed the GNN model architecture of *CG*_*Signature*_, which includes four computational units, each with two-layer *graph convolution* plus one-layer *graph pooling* followed by three-layer fully connected layers (MLP), before generating the prediction output (Figure 1).

The graph convolutional layer is responsible for extracting an array of features from the last output array, which mimics the role of CNN convolution. It changes the dimension *d* of the feature array but does not change the number of nodes *N*_*i*_. The output of graph convolutional layers is passed on to the graph pooling which compresses the node number by a fractional proportion while in this process usually the key structural information and node features are preserved. The MLP readout will then output the label class.

Graph convolution communicates the structural information of the data to the deep network model via the message passing between the neighborhood nodes, which contributes as the key to successfully capturing the geometric feature of the data. In this work, we adopted the GINConv^21^ as the graph convolution and TopKPool^20^ as the graph pooling method, respectively. The convolutional layer for GIN can be aggregated by

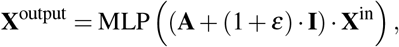

where **X**^in^ ∈ ℝ^*N*×*d*^ is the *d*-feature matrix on the nodes of the graph with *N* nodes for the input layer, and *A* ∈ℝ^*N*×*N*^ is the adjacency matrix of the graph. *W* is the filter weight parameter matrix with the size of *m* × *n* to be learned by the GNNs, where *n* is the number of hidden neurons. GINConv is a special neural message passing operator for GNN aggregation.

Our GNN model was trained by connecting multiple layers of graph convolution activated by a ReLU (Rectifier Linear Unit)^37^. The graph pooling, which is used between two consecutive layers, serves to reduce the dimensionality of the feature map so that the network has appropriate amounts of parameters to circumvent over-fitting^38^. Here we used *TopKPooling*^20^ for graph pooling.

There exist different types of GNN models in the ma-. chine learning literature^39^. Specifically, we tested the performance of the GINConv+TopKPool model with the other three popular GNN models, i.e. GINConv+SAGPool, GCNConv+TopKPool, and GCNConv+SAGPool. The results showed that the chosen model (GINConv+TopKPool) achieved the highest AUROC value and stable training performance. Refer to Figures 2a and 2b for a detailed illustration of the results.

### Hyperparameter optimization

We fine tuned the hyperparameters for the GNN models with the assistance of HyperOPT^25^ and Ray^26^, where the network architecture and batch size were fixed. The hyperparameters were searched within the range as shown in Table 4. More specifically, the best-performing model used the following hyperparameters: learning rate 5 × 10^−4^, weight decay rate 10^−4^, number of hidden neurons 512, pooling ratio 0.5, number of hidden layers 4, batch size 256, and maximal number of epochs 200 with the early stopping strategy.

**Table 4.** Search space for hyperparameters of GNN models.

| Hyperparameter | Searching space |
| --- | --- |
| Learning rate | $10^{-4}, 5 \times 10^{-4}, 10^{-3}$ |
| Weight decay ($L_2$) | $10^{-4}, 5 \times 10^{-4}, 10^{-3}$ |
| Hidden units | 256, 512 |
| Pooling ratio | 0.5, 0.65, 0.75 |

### Prediction aggregation to assess the patient-level performance

The model performance was evaluated at the Cell-Graph level. After the model was optimized, the patient-level performance of the model was calculated by aggregating the prediction results produced by the optimized model. In particular, we fed Cell-Graphs of the test dataset to the optimal model to predict the label for each of them. Since hundreds of Cell-Graphs were sub-sampled from the mIHC images of the patient, hundreds of the predictions were also made for a given patient. To generate the patient-level prediction for a patient, we calculated the proportion of Cell-Graphs belonging to a specific class, and then classified the patient as the group that received the largest proportion of the Cell-Graphs.

### Framelet analysis to facilitate interpretation of the model prediction

From the mathematical perspective, the *framelet system*^31–33,35^ refers to a set of functions that provide a multi-scale representation of graph structured data, which has a similar property to the traditional wavelet in the Euclidean space. Using the framelet transforms, we can decompose the graph features into low-pass and high-pass frequencies as the extracted features to train network models, via the framelet-based graph convolution.

Suppose 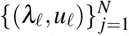 are the pairs of the eigenvalue and eigenvector for the graph Laplacian *ℒ* of a graph *𝒢* with *N* nodes. The (undecimated) framelets at the *scale level j* = 1, …, *J* for graph *𝒢* with the above scaling functions can be defined, for *n* = 1, …, *r*, as follows:

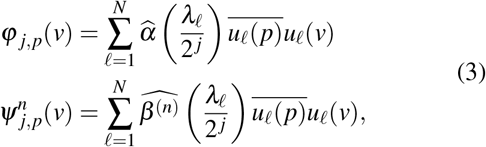

where *φ* _*j,p*_ and 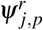 are the low-pass and high-pass framelets translated at the graph node *p*. In the framelet analysis above, we have shown the low-pass and high-pass *framelet coefficients v* _*j,p*_ and 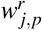 for a signal *f* on graph *𝒢*. They are the projections ⟨*φ* _*j,p*_ *f* ⟩ and 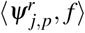 of the graph signal onto framelets at the scale *j* and node *p*. The construction of framelet system and the framelet transforms rely on the filter bank (a collection of filters) to calculate framelet coefficients. Here we used the filter bank of the Haar-type filters for the experiments.^31,35^. The dilation factor is 2 ^*j*^ with the *dilation* (base) 2 for a natural number *j*, where *j* indicates the scale level and 2 ^*j*^ is the scale of the framelet. A bigger value of *j* indicates that the corresponding framelet coefficient carries more detailed information of the graph signal.

The above framelet system is a *tight frame*, which provides an exact representation of any *L*_2_ function on the graph. This guarantees that the framelet coefficients have a unique representation of a graph signal. Accordingly, the framelet coefficients can fully reflect the feature of the signal. Moreover, the coefficients decompose the signal at multi scales and can be used to observe whether a particular scale, or the high-pass or low-pass frequencies contain a more important feature of the data.

## Data Availability

The data used for the main analyses presented here is available for non-commercial use and can be accessible by request.

## Ethics declaration

This study was approved by the Shanghai Ruijin Hospital under protocol 2021SQ015. All researchers were blinded to the patient private data during the experimental analysis.

## Code availability

All the related scripts and code are publicly available and can be download at https://github.com/docurdt/Cell-Graph_Signature.git.

## Acknowledgements

The authors would like to thank the Ruijin Hospital affiliated with Shanghai Jiao Tong University School of Medicine for providing the support of this project. We would also like to thank all the collaborators and colleagues for the enlightening discussions and feedback. This work was supported by the Major Inter-Disciplinary Research (IDR) Grant awarded by Monash University.

## Author contributions

Y.W. and Y.G.W. conceived and conducted the experiments, analysed the results, and wrote the first draft. Y.W., Y.G.W., C.H., M.L., P.L., G.I.W., and J.S. were responsible for the methodology and experiment design. Y.F., N.O., T.K., R.J.D., J.Z., A.B. and G.M. helped to analyse the results. I.S., Q.G., Y.H., and D.X. processed and curated the mIHC data. P.L., D.X., G.I.W., and J.S. supervised this study. All authors reviewed or revised the manuscript.

## Additional information

### Hardware

All the experiments were performed using PyTorch Geometric^40^ on a server with Intel(R) Core(TM) i9-9820X 230 CPU 3.30GHz, NVIDIA GeForce RTX 2080Ti and NVIDIA TITAN V GV100.

